# *NFIX* missense variants that disrupt the β-hairpin loop result in a severe form of Malan syndrome in adolescence with rapidly evolving scoliosis and muscle wasting

**DOI:** 10.64898/2026.07.16.26357549

**Authors:** Christal G. Delagrammatikas, Louise J. Gourlay, Manuela Priolo, Rosaria Russo, Amirsadra Ahmadi, Alberto Giuseppe Barbiroli, Riccardo Capelli, Kaitlynn Stowers, Olivia D’Annibale, Matthew Ravalin, Marco Tartaglia, Marco Nardini, Benjamin T. Cocanougher

## Abstract

**Purpose:** Pathogenic variants in *NFIX* cause Marshall-Smith syndrome and Malan syndrome (MALNS). We identified a severe subtype of MALNS characterized by adolescent-onset musculoskeletal deterioration and investigated functional consequences of underlying variants.

**Methods:** Clinical data were collected from seven individuals with pathogenic *NFIX* variants. Wild-type and mutated recombinant NFIX DNA-binding domains (DBDs) were evaluated using biochemical, structural, and DNA-binding assays.

**Results:** Six individuals carrying R116W, R116P, K125E, or G147E NFIX substitutions developed progressive muscle wasting, markedly reduced body mass index, and rapidly progressive scoliosis after the typical childhood features of MALNS; two died from disease-related complications. A seventh individual with R116G did not develop this severe phenotype. Functional studies on recombinant NFIX DBDs showed complete or near-complete loss of DNA-binding activity for R116W, R116P, K125E, and G147E despite preserved protein folding, consistent with disrupted DNA recognition and a potential dominant-negative mechanism. In contrast, R116G exhibited a 7.7°C decrease in thermal stability, which may support haploinsufficiency mediated by protein degradation.

**Conclusion:** Specific *NFIX* missense variants define a severe subtype of MALNS associated with progressive musculoskeletal deterioration. *In vitro* functional studies support variant-specific disruption of DNA binding, providing a mechanistic basis of genotype-phenotype correlations and informing prognosis, clinical surveillance, and therapy development.

## Introduction

Nuclear factor I X (NFIX) is a member of the NFI family, a ubiquitous group of transcription factors (TFs), encoded by four closely related genes (*NFIA, NFIB, NFIC, and NFIX*), that play essential roles in regulating gene expression, chromatin organization, and cell signaling.^1-4^ *NFIX*-related disorders encompass at least two clinically and genetically distinct disorders, Marshall-Smith syndrome (MRSHSS, MIM: 602535) and Malan syndrome (MALNS, MIM: 614753).^5-9^ MRSHSS is caused by heterozygous frameshift variants within exons 6-10 that escape nonsense-mediated decay (NMD), resulting in a protein with a distinctive C-terminus that retains DNA-binding ability but exhibits aberrant functionality due to its altered transcriptional activation domain (TAD).^6,8^ MRSHSS is characterized by severe skeletal dysplasia, respiratory insufficiency, failure to thrive, and high infant mortality.^7,8^ In contrast, MALNS is associated with heterozygous structural rearrangements partially or totally encompassing *NFIX* or intragenic variants including missense and truncating variants primarily affecting exon 2, which encodes the DNA-binding domain (DBD).

The prevailing model is that MALNS is caused by *NFIX* haploinsufficiency.^6,9-11^ Some missense variants are presumed to reduce protein stability and mediate accelerated protein degradation by the proteasome, furthermore supporting haploinsufficiency.^12,13^ MALNS affected individuals typically present with excessive skeletal growth during childhood, macrocephaly, and developmental delay (DD)/intellectual disability (ID), but are characterized by a comparatively stable clinical course without the severe early-life respiratory or musculoskeletal complications that are typical of MRSHSS.^7,14-16^

Here, we report a previously unappreciated severe presentation of MALNS in six individuals harboring specific *NFIX* missense variants in exon 2 (R116W, R116P, K125E, and G147E) characterized by overgrowth and DD/ID in early childhood with progressive musculoskeletal deterioration in adolescence. We perform biochemical and structural analyses of these variants to explore potential mechanisms which may contribute to disease severity. These findings expand the phenotypic spectrum of *NFIX*-related disorders and suggest the importance of variant-level functional characterization for elucidating genotype-phenotype relationships.

## Methods

### Ethics statement and Patient Ascertainment

Informed consent was obtained for release of medical information and photographs under CCHMC IRB 2025-0631 (Rare Genetic Disease Biorepository) and 2025-0338 (Muscle Wasting in Malan Syndrome). Participants were recruited through the MALNS patient registry (Sanford CoRDS) and direct emails from the patient advocacy group. Inclusion criteria included: 1) molecularly confirmed MALNS clinical diagnosis, 2) musculoskeletal involvement assessed by body mass index (BMI) and spine curvature, and 3) willingness to share medical history and genetic test results. Medical records and caregiver-reported data were obtained and stored securely in a Research Electronic Data Capture (REDCap) database. All individuals exhibiting a severe musculoskeletal phenotype were included in the study. Individuals with changes at the same codon and presenting with a less severe phenotype were not excluded. Participant 7 was included as he shared a different missense change at codon 116, though not presenting with low BMI and progressive severe scoliosis.

### Protein variant production and DNA binding assays

MALNS-associated variants (R116W, R116P, R116G, K125E, and G147E) were introduced in the NFIX DBD coding sequence by site-directed mutagenesis, using the wild-type NFIX DBD plasmid as a template, and Phusion DNA polymerase (Thermofisher Scientific), following the manufacturer’s instructions. All constructs were expressed in Shuffle(DE3) bacterial cells (New England Biolabs) and products were purified, as previously described.^12^ Electrophoretic mobility shift assays (EMSA) were performed as previously reported using a 6% native polyacrylamide gel and a Cy5-labeled 31 bp dsDNA probe containing the NFI consensus sequence.^12^

## Results

We systematically assessed the clinical presentation of six participants with MALNS and severe musculoskeletal deterioration (Figure 1; Table S1). Structured interviews and telehealth evaluations were performed with parents/caregivers to more accurately define the natural history of this globally distributed cohort. The interviews revealed a typical MALNS developmental trajectory characterized by overgrowth and DD in early childhood. Most individuals experienced neonatal feeding difficulties, congenital brain anomalies, or congenital cardiac defects, within the range expected for MALNS. During childhood, most individuals maintained a tall, slender habitus despite preserved caloric intake.

**Figure 1.**
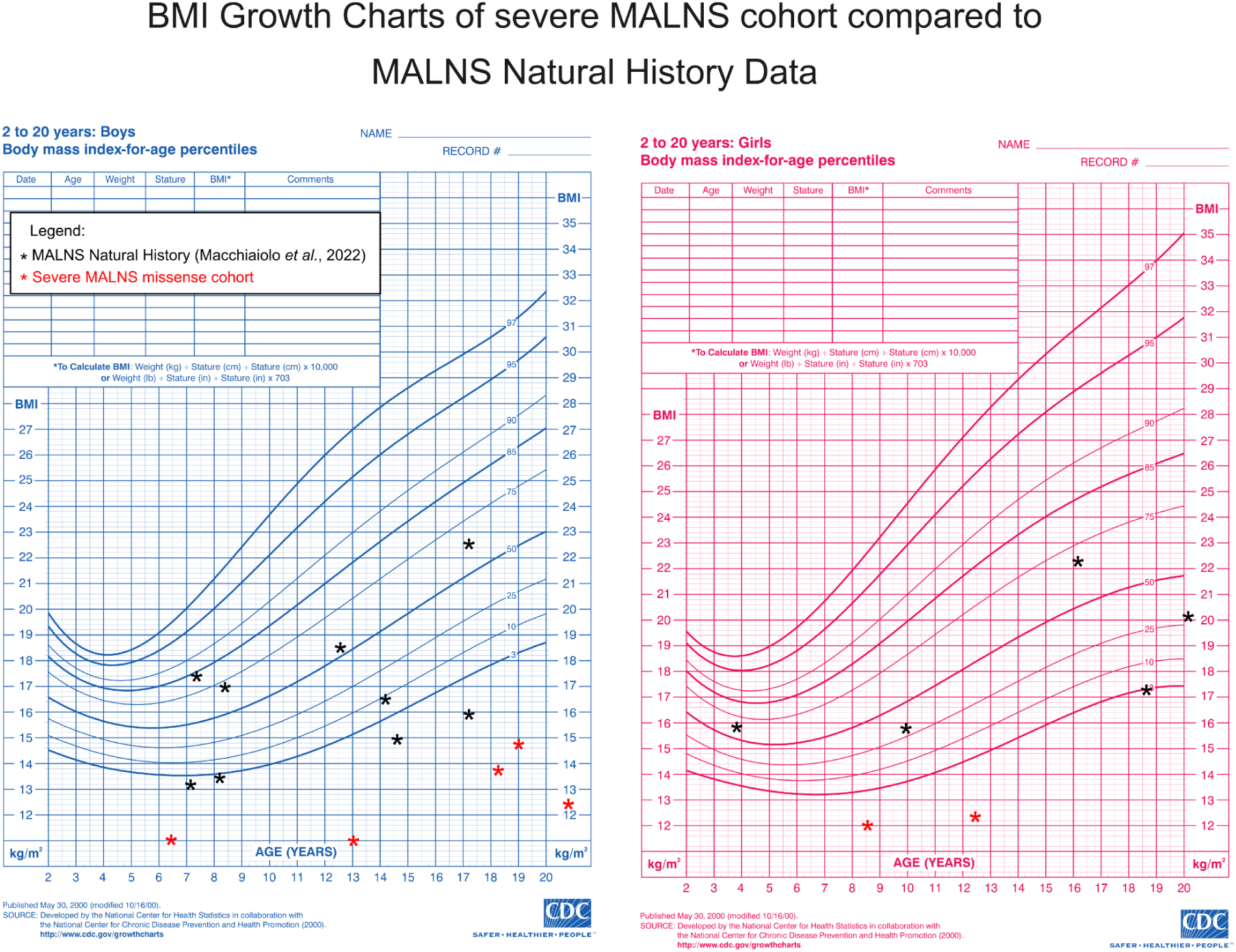
Progressive musculoskeletal deterioration associated with specific NFIX missense variants in Malan syndrome. BMI-for-age growth charts comparing individuals with severe NFIX missense variants (red symbols) with published MALNS natural history data^15^ (black symbols). (Clinical photographs have been removed from the preprint version to preserve participant anonymity. Researchers may contact the corresponding author regarding access to these materials).

A strikingly consistent change in disease course emerged in late childhood (participants 3, 4, and 5) or early adolescence (participants 1, 2, and 6). Participants 1-6 developed progressive loss of muscle mass accompanied by declining BMI (Z score < -3) despite adequate nutritional intake, followed by rapidly progressive scoliosis (participants 1, 2, 4 and 6), reduced exercise tolerance, and progressive motor impairment. Two individuals developed severe cardiac and respiratory compromise secondary to scoliosis and died during adolescence or early adulthood.

Participants in this clinically identified cohort were found to be heterozygous for *NFIX* missense variants resulting in one of the following substitutions: p.R116W, p.R116P, p.K125E, or p.G147E. Given the similar location of these amino acid changes in the protein structure, we asked whether individuals with other substitutions at the same residues may also exhibit a severe phenotype. An individual with an R116G change was identified for comparison (Figure 1F, Table S1, participant 7). This individual exhibits the typical MALNS neurodevelopmental phenotype, but in adolescence, he has preserved muscle mass and a BMI in the normal range. Based on these observations, we proceeded with functional testing of these missense variants to investigate their impact on NFIX structure and function.

### MALNS-associated mutant DBD variants show impaired DNA binding

Wild-type and mutant (R116W, R116G, R116P, K125E, and G147E) NFIX DBDs were produced in bacterial cells, as previously described^12^ (Figure S1a), with significantly reduced yields for all mutants compared to the wild-type DBD (Table S2). These data suggest a reduced folding efficiency. Size exclusion chromatography (SEC) showed that the R116W, R116G, K125E, and G147E NFIX DBDs were monomeric in solution, like the corresponding wild-type domain (Table S2), indicating that the mutations do not influence the oligomeric state of the proteins.

To evaluate the impact of variants on protein folding and stability, circular dichroism (CD) and thermal stability analyses were performed on the R116G, R116W, K125E and G147E mutants.^12^ Similar analyses could not be performed for R116P due to low protein yield. Thermal stability was comparable to wild-type DBD, except for R116G, which was 7.7°C less stable, and R116W, which showed a 3.4°C reduction in stability (Table S2; Figure S1b). The relative stabilities of R116W and R116G may contribute to the distinction in phenotypes for these pathogenic variants.

*In silico* molecular dynamics (MD) analyses were performed on the K125E, G147E, and R116W DBDs, the latter being the most phenotypically severe R116 substitution. MD simulations showed no major changes in secondary structure or overall DBD fold stability (Figures S2 and S3). G147E displayed minor structural deviations, with slight destabilization of the C-terminal region (Figure S2).

DNA binding activity was assessed by EMSA. Within the NFIX DBD, R116 and K125 are the only residues that establish base-specific contacts with DNA (Figure 2a). In the consensus sequence (T^9^T^10^G^11^G^12^C^13^), R116 interacts with G^12^ and G^13^ on the complementary strand, whereas K125 contacts G^11^ and G^12^.^12^ Consistent with their critical role in sequence recognition, substitution of R116 with a glycine (no side-chain), proline (geometrically-restricted cyclic side chain), or substitution of K125 with a negative charge (K125E), completely abolished DNA binding activities (Figure 2b for R116G, and previously reported^12^ for R116P and K125E). DNA-binding was also absent for the R116W variant, although at high protein concentrations (8-fold relative to the DNA probe), a faint smear was observed in EMSA, indicating a residual, albeit unstable, capacity for DNA interaction (Figure 2b).

**Figure 2.**
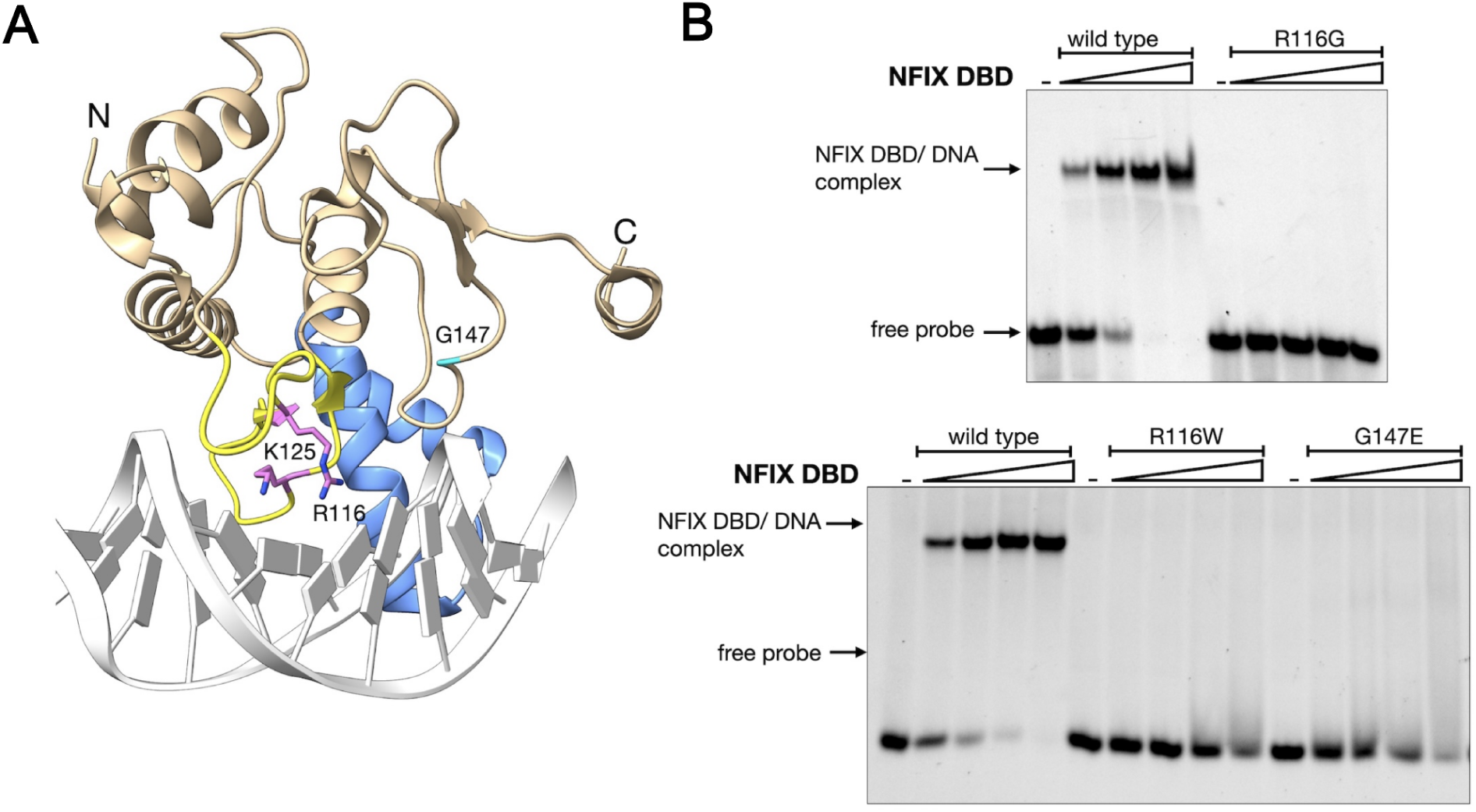
Mapping MALNS-causing variants on the structure of the NFIX DNA-binding domain. **A)** The three affected residues are mapped on the three-dimensional structure of a single NFIX DBD monomer (ribbons) bound to a NFI half site in double-stranded DNA (white cartoon) (PDB: 9QKY^12^). The side-chains of R116 and K125 are shown as pink sticks, whereas G147, is highlighted in cyan. The NFI-specific antenna domain and the MH1-like core domain are colored blue and tan, respectively, and the β-hairpin loop, critical for DNA-binding, is shown in yellow. The N- and C-termini are labeled and frontal clipping was used for clearer visualization. This panel was generated using UCSF ChimeraX version 1.9; **B)** DNA-binding ability was assessed by dose-response EMSA experiments, carried out with wild type NFIX DBD and variants R116G, R116W and G147E; EMSA results for R116P and K125E were previously reported.^12^ DNA binding abilities were measured using a Cy5-labeled 31 bp dsDNA probe, containing the NFI consensus sequence, as detailed in Tiberi et. al., 2025. Briefly, 20 nM dsDNA probe was incubated in the absence (-) or presence of increasing protein concentrations (X1, X2, X4, X8 with respect to the probe). Bands corresponding to the free dsDNA probe or the NFIX DBD/DNA complex are indicated.

In contrast, G147 does not directly participate in DNA binding (Figure 2; Figure S4),^12^ but stabilizes the β-hairpin that inserts into the major groove for DNA recognition by forming a hydrogen bond (3.2 Å) with the side-chain of β-hairpin residue R128 (Figure S4). Pathogenic variants at residue R128 have been reported in MALNS.^9^ Substitution of G147 with a glutamate places it in proximity to another glutamate, E148, likely resulting in electrostatic repulsion and local structural perturbation. In addition, MD studies revealed that E147 rapidly forms a salt bridge with the neighboring R116 side chain, sequestering the β-hairpin and preventing correct DNA interactions (Figure S5). DNA binding was largely abolished for G147E, although at high protein concentrations (8-fold relative to the DNA probe) a weak smeared band was observed in EMSA, indicative of unstable complex formation (Figure 2b).

## Discussion

Here, we identify a previously unappreciated severe manifestation of MALNS characterized by adolescent-onset muscle wasting and rapidly evolving scoliosis, resulting in a fatal course in two of six affected individuals. This phenotype was observed in subjects with variants altering either one of the ◻-hairpin DNA contact residues R116 and K125 or the β-hairpin stabilizing residue G147.^12^ The milder phenotype observed in individuals with R116G (participant 7 here and previous reports^11,15^), compared to the more severe phenotype in individuals with R116W (participants 1 and 2 and a previous report^17^), suggests that a different substitution at the same residue can influence disease severity. Of note, a female individual carrying the R116P substitution, previously reported in childhood,^15^ did not present with low BMI, in contrast to the age-matched male individual (participant 3) in the present cohort, suggesting that genetic or biological modifiers, including sex, may contribute to disease presentation.

Functional studies performed here demonstrate profound impairment of DNA binding despite preservation of overall protein folding, indicating that these variants disrupt DNA recognition rather than causing global destabilization of the DNA-binding domain. The disease mechanism remains uncertain, but it may be compatible with either dominant-negative effects through nonproductive dimer formation or haploinsufficiency from impaired protein stability.

A plausible dominant-negative model is that mutant proteins participate in non-productive dimer formation. Given that NFIX can bind NFI half-sites with reduced affinity^18^ as a monomer but also as dimers, albeit unstable^12^, a wild-type monomer may engage DNA and subsequently recruit a mutant partner, generating a transcriptionally inactive heterodimer. Such a mechanism would be consistent with a dominant-negative effect and provides a potential explanation for the severe phenotypes observed. In parallel, the reduced yield of recombinant variants in *E. coli* suggests impaired folding efficiency, likely leading to enhanced proteolytic clearance. If recapitulated in eukaryotic systems, the combination of reduced protein levels and defective DNA binding would support a haploinsufficiency mechanism. These mechanisms are not mutually exclusive and may act in concert in a variant- and context-dependent manner. Consistent with this, such an articulated effect is characteristic of disorders involving proteins that assemble into functional complexes, where concomitant defects in assembly and stability synergistically exacerbate functional loss.

These findings have important clinical implications. Experimental studies demonstrate that NFIX represses myostatin expression, providing a biologically plausible mechanism linking NFIX dysfunction to progressive muscle wasting.^19,20^ Although causality remains to be established in MALNS, these observations support evaluation of early musculoskeletal surveillance, timely scoliosis management, and future investigation of clinically advanced myostatin inhibitors for affected individuals.

## Data Availability

All clinical information is included in the main text. Supplemental Table 1 the Supplemental Case Reports have been removed from the preprint version to protect participant anonymity. If further details are required, data will be supplied upon reasonable request by qualified investigators for follow-up research studies or personalized patient care. Please send requests to.

## Acknowledgments

The authors gratefully acknowledge the individuals with Malan syndrome and their families who participated in this study. Their generosity in contributing clinical information and their ongoing commitment to research are fundamental to advancing our understanding of Malan syndrome and improving care for affected individuals. The authors also thank the Malan Syndrome Foundation for its invaluable partnership and steadfast support of the Malan syndrome research community.

## Funding Statement

BTC received funding through the Cincinnati Children’s Research Foundation to support this work.

## Author Contributions

Conceptualization: C.G.D., L.J.G., M.N., B.T.C.; Data curation: C.G.D., L.J.G., M.P., M.T., M.N., B.T.C.; Formal analysis: C.G.D., L.J.G., M.P., M.T., M.N., B.T.C.; Funding acquisition: M.N., B.T.C.; Investigation: C.G.D., L.J.G., M.P., R.R., A.A., A.G.B., R.C., K.S., O.D., M.R., M.T., M.N., B.T.C.; Methodology: C.G.D., L.J.G., M.N., B.T.C.; Project administration: M.N., B.T.C.; Resources: M.P., M.T., M.N., B.T.C.; Supervision: M.N., B.T.C.; Visualization: L.J.G., B.T.C.; Writing-original draft: C.G.D., L.J.G., M.N., B.T.C.; Writing-review & editing: C.G.D., L.J.G., M.P., M.R., M.T., M.N., B.T.C.

## Ethics Declaration

This study was approved by the institutional review board at Cincinnati Children’s Hospital Medical Center (IRB #2025-0631 and #2025-0338). Legal guardians provided written informed consent for study participation, data review, and photo release where applicable, which has been received and archived by the study team.

## Conflict of Interest

The authors declare no conflicts of interest related to this work.

## Declaration of generative AI and AI-assisted technologies in the manuscript preparation process

During manuscript preparation, BTC used ChatGPT (OpenAI) to improve readability, identify opportunities to reduce redundancy, and suggest ways to condense the text. The AI tool was not used to generate scientific content, analyze data, interpret results, or develop conclusions. All suggestions were critically reviewed, edited, and verified by the authors, who take full responsibility for the content of the manuscript.

## Notes

### Competing Interest Statement

The authors have declared no competing interest.

### Author Declarations

The IRB of Cincinnati Children's Hospital gave ethical approval for this work.

