## Supplemental_Figures for "*NFIX* missense variants that disrupt the β-hairpin loop result in a severe form of Malan syndrome in adolescence with rapidly evolving scoliosis and muscle wasting"

**Supplementary Figures**

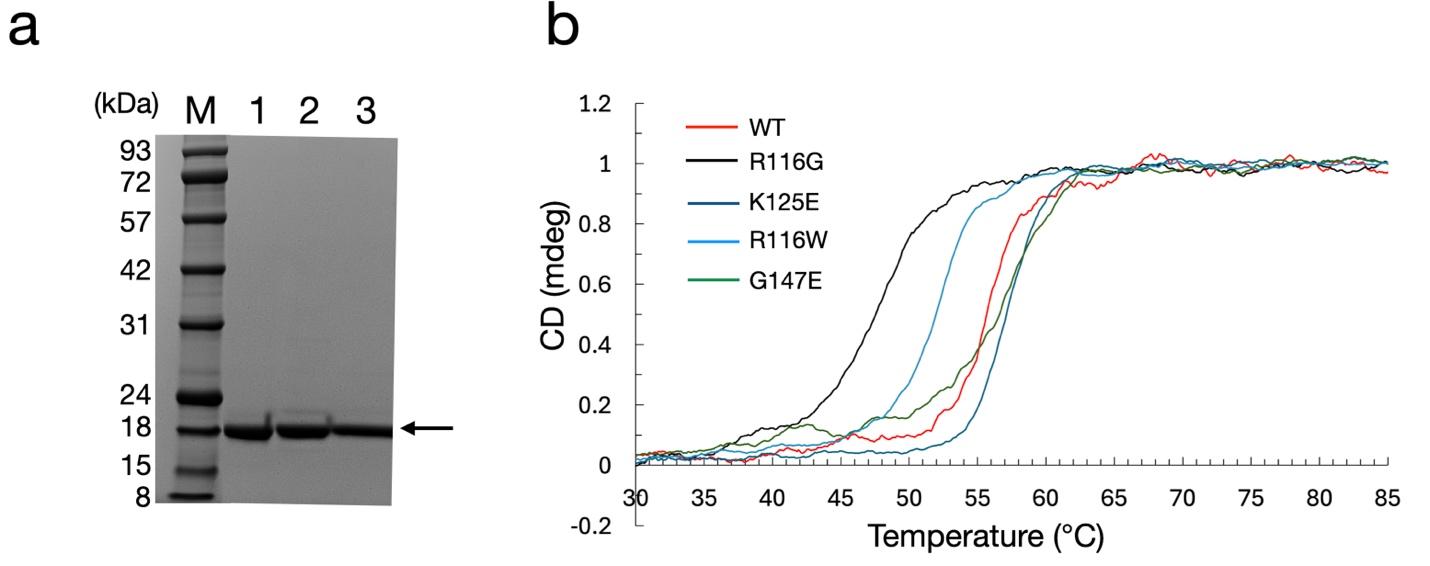

**Figure S1. Biochemical and biophysical characterisation of NFI-X constructs. a)** SDS-PAGE analysis of purified R116W (lane 1), K125E (lane 2) and G147E (lane 3) samples on a 12% mPAGE™ 12% Bis-Tris Precast Gel (GeneScript Biotech). Data are not shown for R116G and R116P and are available upon request. All mutants migrate with a MW around 21 kDa, in line with their calculated molecular weights (MWs) and as for wild type DBD (Tiberi et al., 2025). The MWs for the StoS Prestained Protein Ladder (GeneSpin) are shown on the left; **b)** the stability of Malan DBD variants was assessed by circular dichroism (CD), in comparison with the wild-type, tracing the variation in ellipticity at 222 nm wavelength that follows the loss in α-helix CD signal contribution, as a function of temperature. CD was not carried out on R116P due to low protein yields. Unfolding curves are expressed as the proportion of total unfolded protein versus temperature (°C). CD profiles were generated using the SigmaPlot v.14 (Systat Software inc.).

**Figure S2**

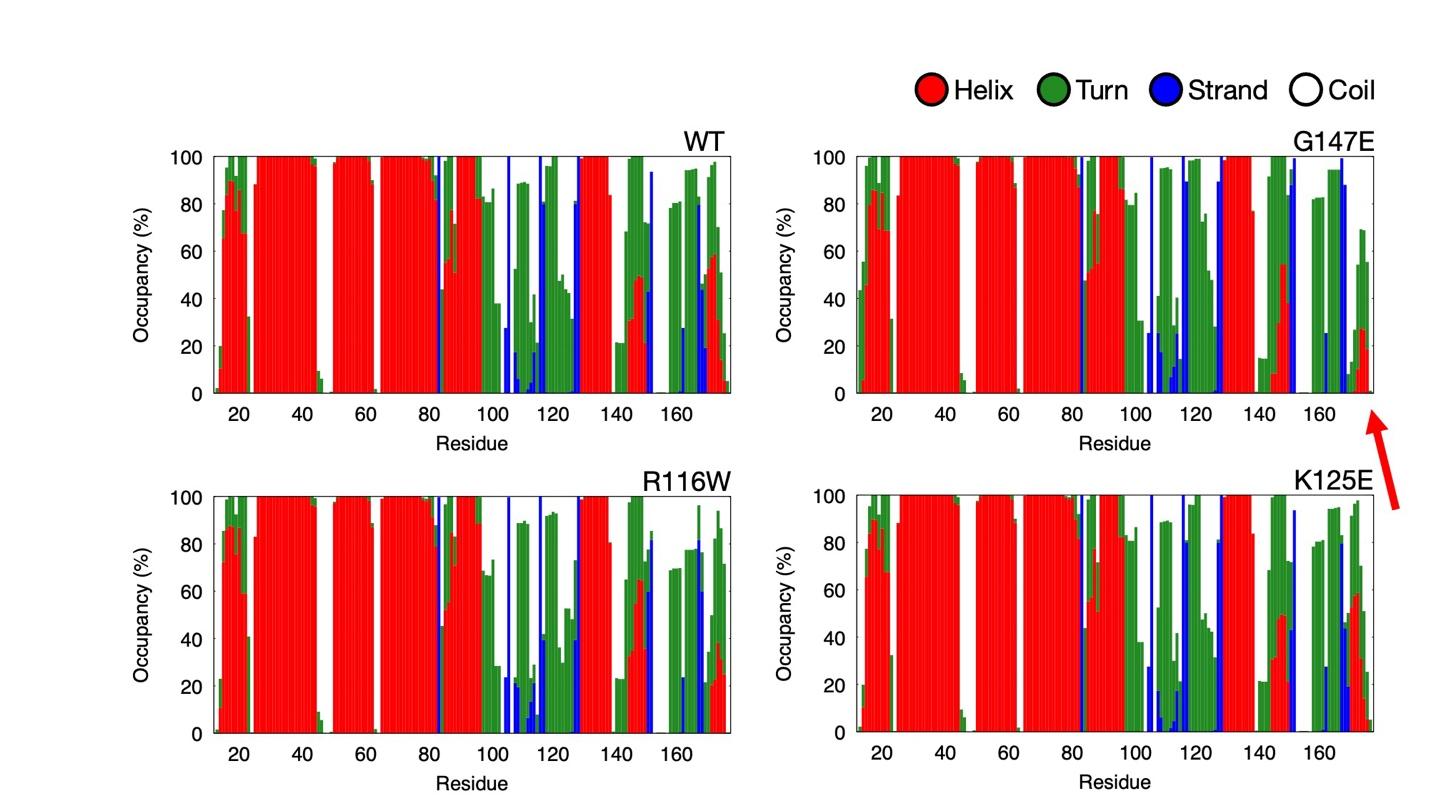

**Figure S2. *In silico* secondary structure analyses of Malan Variants.** We monitored the secondary structure of NFIX DBD monomer (residues 13-178) for wildtype and three Malan variants that lead to the severe patient phenotype (R116W, G147E, and K125E) using the stride software. No evident secondary structure deviations were observed for different mutants, except for G174E, which resulted in a statistically significant lower probability to have an α-helix at residues 171-177.

**Figure S3**

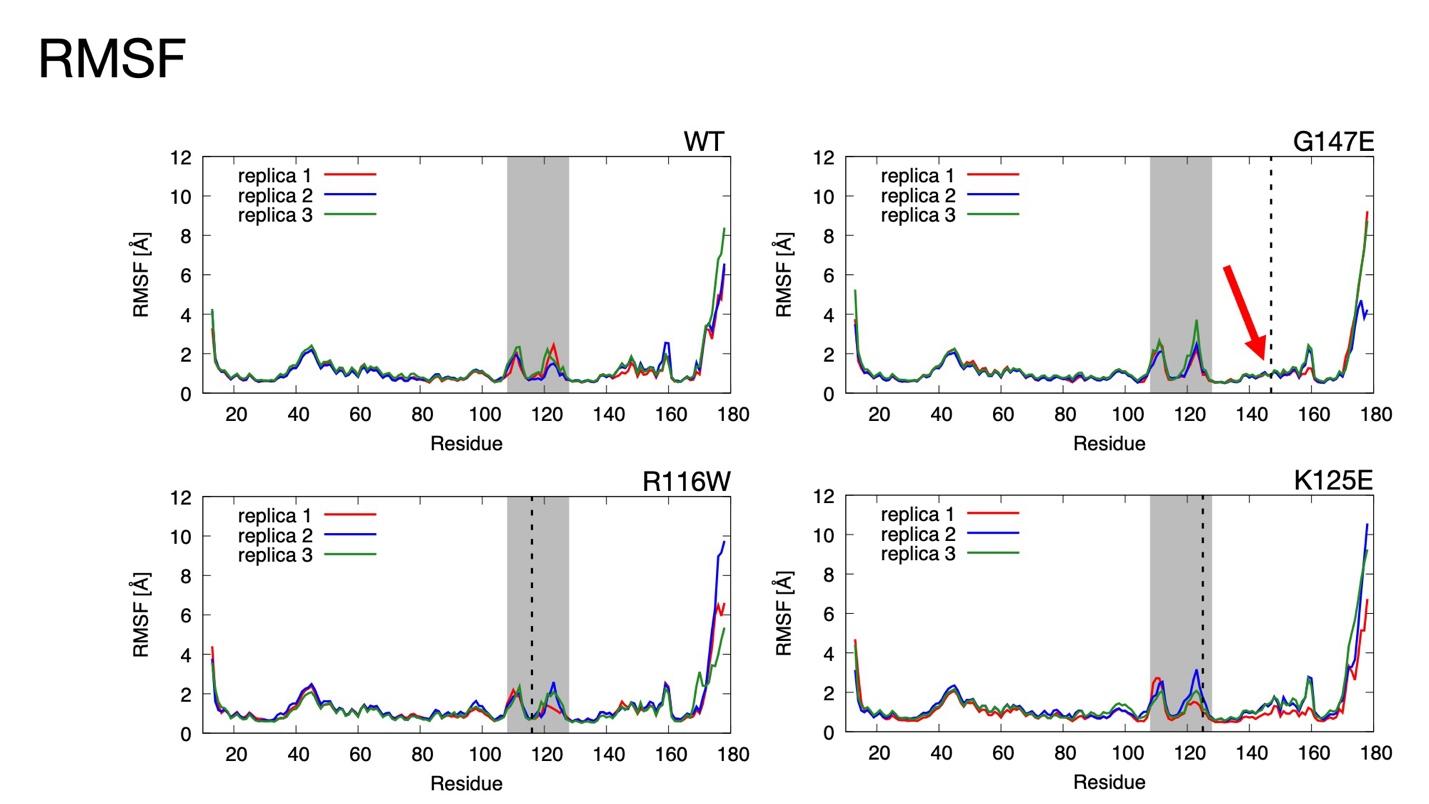

**Figure S3. Root mean square fluctuation for WT NFIX and mutants.** We computed the per-residue RMSF of wild type, R116W, K125E and G147E models, highlighting the DNA-binding loop in grey and the mutation site with a dashed line. The only difference that was observed among the four different RMSF profiles was the slightly more stable behaviour of the region corresponding to residues 145-155 in G147E (highlighted by a red arrow).

**Figure S4**

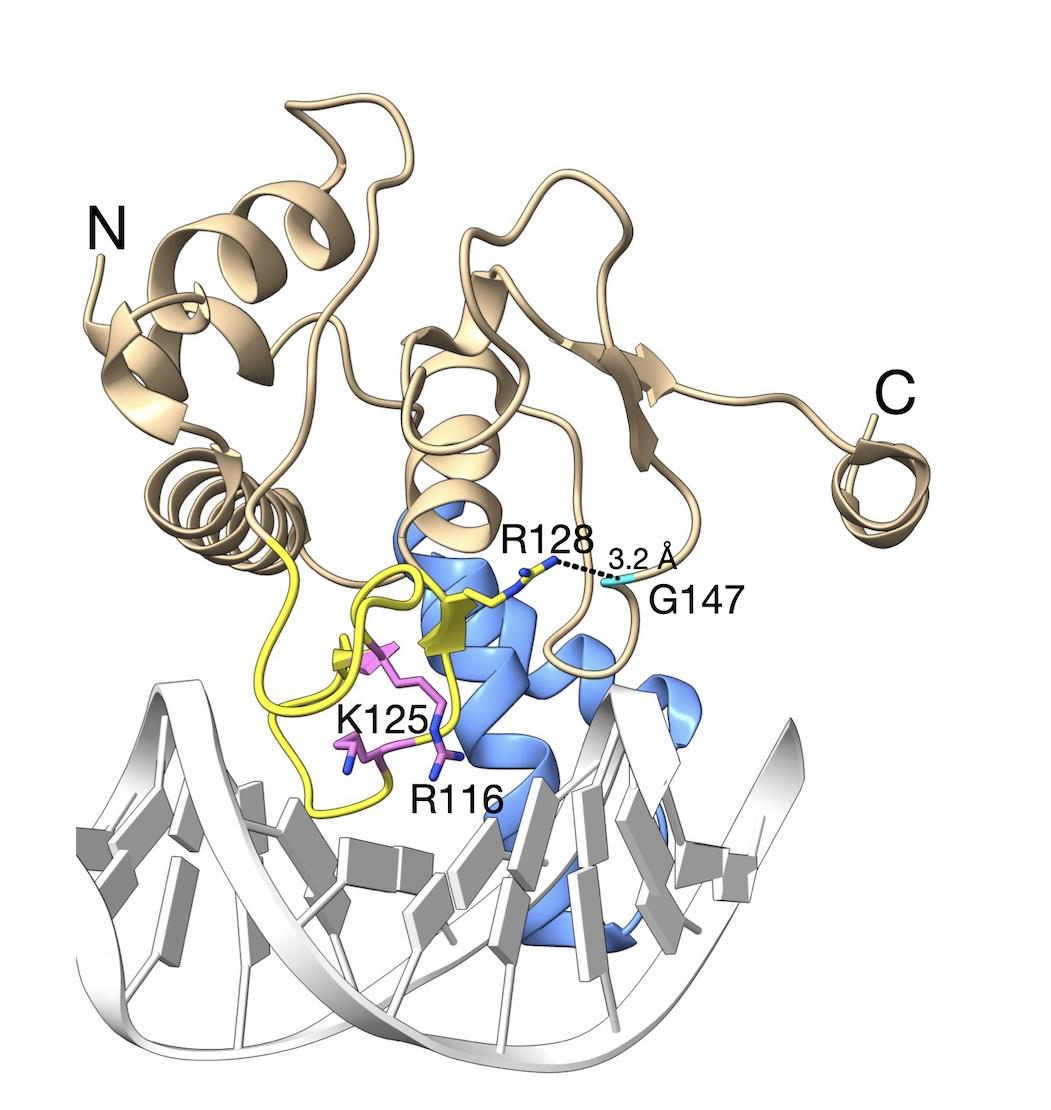

**Figure S4. Structural analysis of G147.** The stabilizing H-bond (3.2 Å) formed between the carbonyl oxygen atom of G147 (cyan) and side chain NH2 atom of β-hairpin residue R128, are shown on the three-dimensional structure of the NFIX^DBD^ monomer (ribbons), bound to a NFI half site in double-stranded DNA (white cartoon) (PDB: 9QKY (Tiberi et al., 2025). The other residues that are the focus of this study are shown as sticks. Coloring and labelling is as described in main text Figure 1. This figure was generated using UCSF ChimeraX version 1.9 (Meng et al., 2023; Petterson et al., 2021).

**Figure S5**

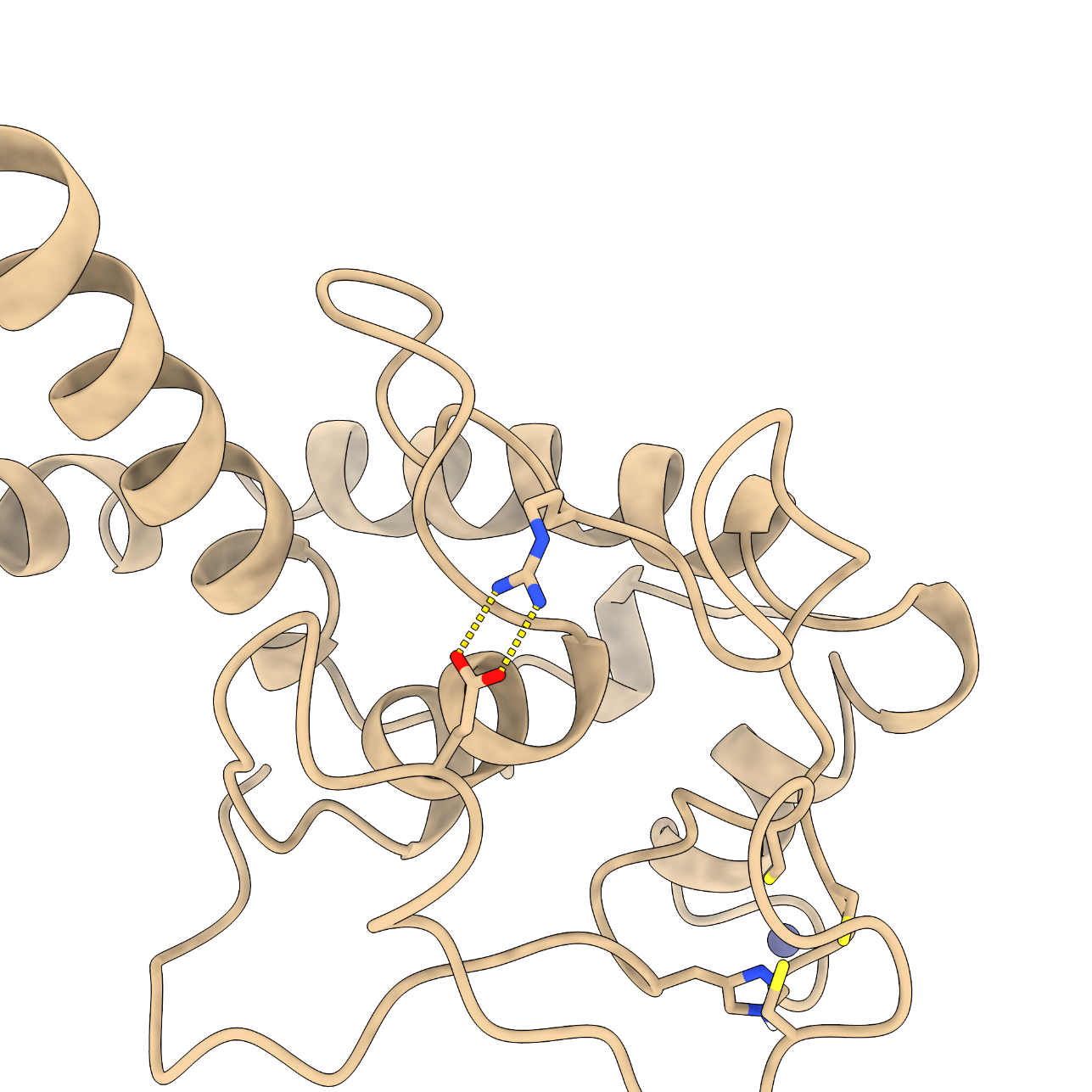

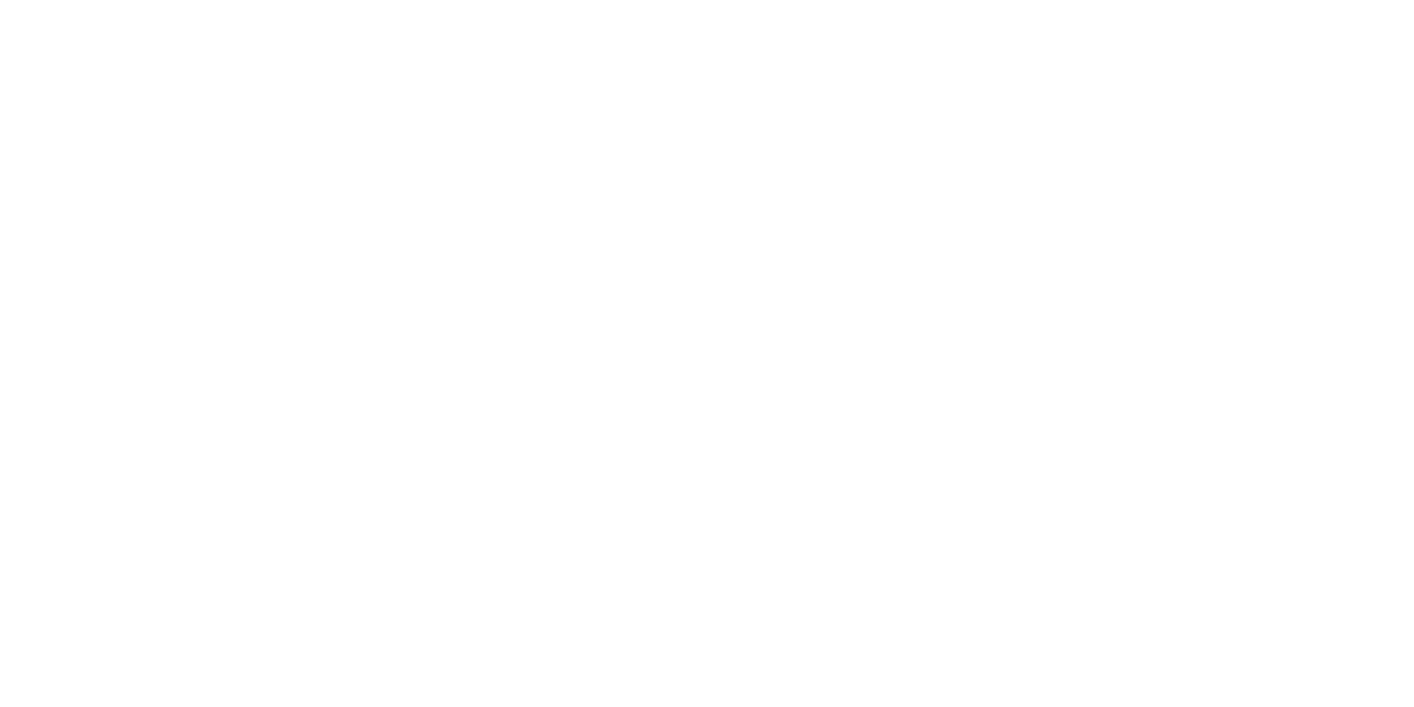

R116

E147

**Figure S5. *In silico* analysis of the effect of E147.** The presence of E147 in the structure of NFIX DBD (beige ribbons) over stabilizes the DNA binding β-hairpin loop by forming a salt bridge with R116 (left panel) that is likely to hinder correct DNA binding. E147 and R116 are shown in sticks and labelled. Oxygen and nitrogen atoms are colored red and blue, respectively. The formation of this non-bonded interaction between E147 and R116 was consistently observed in all three replicates (right panel).

**Table S2**

| **DBD**  **Variant** | **Expression yield (mg/L culture)** | **SEC Elution Volume (V_e_)**  **(mL)** | **T_M_ value**  **(°C)** |
| --- | --- | --- | --- |
| wild type  R116G  R116P  R116W  K125E  G147E | 2.10  0.2  0.05  0.9  0.6  0.1 | 18.5  18.6  -  18.3  18.2  18.2 | 55.7  48.0  -  52.3  57.2  57.4 |

**Table S2: Expression yields, oligomeric state and thermal stability of Malan mutant DBDs.** Purified recombinant protein yields (mg) of Malan variant NFIX DBDs per litre of bacterial culture, produced, following the protocol described in Tiberi et al., 2025, are shown in comparison with wild type NFIX DBD. Oligomeric states of protein variants (except R116P, due to insufficient sample yields), in comparison with wild type, was assessed by size exclusion chromatography (SEC), as previously described (Tiberi et al., 2025). Elution volumes (mL) are shown for all variants and are indicative of monomeric states in solution. Thermal stabilities of all variants, except R116P, were estimated from melting temperature (T_M_) values, measured using circular dichroism (CD), following the loss α-helix CD signal contribution (measured at 222 nm wavelength), as a function of temperature. The inflection points of unfolding curves, generated as described in Figure S1, were derived by taking the first derivative of the data, thus representing the melting temperature.

**Supplementary Methods**

**Molecular Dynamics simulations**

Molecular models of NFIX DBD and its Malan variants were generated starting from the crystal structure with PDB ID 7QQD (Tiberi et al., 2025). The nucleic acid and the second protein chain were removed from the original structure, retaining a single NFIX monomer. R116W, K125E and G147E variants were generated using the Mutagenesis Wizard in PyMOL. Each system was solvated in an explicit water box of 9 × 9 × 9.5 nm^3^ and neutralized with NaCl at a concentration of 150 mM using AmberTools (Case et al. 2023). The systems were described using the AMBER14SB force field (Maier et al., 2015) for the protein, the TIP3P model (Jorgensen et al. 1983) for water, Joung-Cheatham parameters (Joung et al., 2008) for non-zinc ions, and ZAFF parameters (Peters et al., 2010) or Zn^2+^ and the coordinating residues. Each model was minimized in three consecutive steps: (i) steepest descent minimization until a force tolerance of 1000 kJ mol^-1^ nm^-1^ was reached, (ii) conjugate-gradient minimization until a force tolerance of 100 kJ mol^-1^ nm^-1^ was reached, and (iii) L-BFGS minimization until a force tolerance of 10 kJ mol^-1^ nm^-1^ was reached. After minimization, a 2 ns restrained equilibration was performed, followed by a 0.5 ns unrestrained relaxation under the same conditions. Production simulations were then carried out for 1 µs in triplicate for each system. All molecular dynamics simulations were performed in the NPT ensemble using the velocity-rescale thermostat (Bussi et al., 2007) at 310 K with a time constant of 1 ps, and the C-rescale barostat (Bernetti et al., 2020) at 1 bar with a compressibility of 4.5 × 10^-5^ bar^-1^ and a time constant of 5 ps. Long-range electrostatics were treated using the Particle Mesh Ewald method (Huang et al., 2012), with a short-range cutoff of 1 nm. Nonbonded interactions were treated using a potential-shift cutoff scheme. All the simulations have been performed using GROMACS 2024.4 (Abraham et al., 2015).
